# Stroke After High-Dose/Adjuvanted Influenza Vaccines in U.S. Older Adults; 2016-2019

**DOI:** 10.1101/2024.01.15.24301178

**Authors:** Yun Lu, Kathryn Matuska, Yuxin Ma, Layo Laniyan, Yoganand Chillarige, Steven A. Anderson, Richard A. Forshee

## Abstract

**Importance:** A recent study from the U.S. Food and Drug Administration investigated the risk of stroke following COVID-19 bivalent and high-dose/adjuvanted influenza vaccines among older adults (individuals ≥65 years) who experienced stroke after vaccination in the 2022-2023 season. The study found a small but significant association between stroke and high-dose/adjuvanted influenza vaccination in the Medicare fee-for-service (FFS) population.

**Objective:** To evaluate stroke risk following high-dose/adjuvanted influenza vaccines in three historical influenza seasons.

**Design, Setting, and Participants:** The analysis included Medicare beneficiaries ≥65 years who had a stroke outcome after receiving a high-dose or adjuvanted influenza vaccine in three influenza seasons 2016-2019. For each season, the study period began on the first Sunday of August and ended one day before the start of the subsequent season (e.g., Sunday, August 7, 2016 through Saturday, August 5, 2017). A self-controlled case series analysis was performed to compare the risk of stroke in risk intervals (1-21 and 22-42 days) to a control interval (43-90 days).

**Exposures:** High-dose/adjuvanted influenza vaccines.

**Main Outcomes and Measures:** Non-hemorrhagic stroke (NHS), transient ischemic attack (TIA), NHS and/or TIA (NHS/TIA), and hemorrhagic stroke (HS).

**Results:** We observed 29,730 stroke cases in 2016-2017; 34,518 cases in 2017-2018; and 36,869 cases in 2018-2019. In 2016-2017, the primary analysis found a statistically significant association for HS during the 22-42 day risk window (incidence rate ratio (IRR)=1.14, 95% confidence interval (CI) 1.02–1.28; Risk Difference (RD)/100,000 doses=0.84, 95% CI 0.14−1.54) compared to the control interval. However, no significant primary analysis results were identified in 2017-2018 or 2018-2019.

**Conclusions and Relevance:** We did not observe clear, consistent evidence of increased stroke risk following high-dose or adjuvanted influenza vaccination across the three seasons 2016-2019. The statistically significant associations we identified were not consistently observed across outcomes, risk windows, age subgroups, or seasons. The clinical significance of any potential risk of stroke following vaccination must be carefully considered together with the significant benefits of receiving an influenza vaccination.

## Introduction

A recent study from the U.S. Food and Drug Administration investigated the risk of stroke following COVID-19 bivalent and high-dose/adjuvanted influenza vaccines among older adults (individuals ≥65 years) who experienced stroke after vaccination.^1^ The self-controlled case series (SCCS) study did not find clear, consistent evidence of a risk of stroke in the days shortly following COVID-19 bivalent vaccination. However, a small but significant association between stroke and high-dose/adjuvanted influenza vaccination was detected in the Medicare fee-for-service (FFS) population in the 2022-2023 season. To determine if an increase in stroke risk occurred in prior years following influenza vaccination in Medicare beneficiaries, we conducted this follow-up historical analysis of the 2016-2017, 2017-2018, and 2018-2019 seasons.

## Methods

Using a SCCS design, the risk of four incident (no events in 365 days prior) stroke outcomes – non-hemorrhagic stroke (NHS), transient ischemic attack (TIA), a combined outcome of NHS and/or TIA (NHS/TIA), and hemorrhagic stroke (HS) – following exposure to a high-dose or adjuvanted influenza vaccine were evaluated, first among persons ≥65 years (primary analysis) and then by age subgroup (65-74, 75-84, 85+ years).^1^ Fixed risk windows (1-21 days and 22-42 days) were compared to a post-vaccination control window (43-90 days). For each season, the study period began on the first Sunday of August and ended one day before the start of the subsequent season (e.g., Sunday, August 7, 2016 through Saturday, August 5, 2017). Our study population consisted of beneficiaries aged ≥65 years who did not reside in a nursing home facility and were continuously enrolled in FFS Medicare plans for at least a year prior to vaccination. We estimated incidence rate ratios (IRR) for each season using conditional Poisson regression and calculated risk difference (RD) per 100,000 doses. We also conducted a retrospective temporal scan to identify clusters of increased stroke risk within 90 days following vaccination.

## Results

We observed 29,730 stroke cases in 2016-2017; 34,518 cases in 2017-2018; and 36,869 cases in 2018-2019 (Table 1). In 2016-2017, the primary analysis found a statistically significant association for HS during the 22-42 day risk window (IRR=1.14, 95% confidence interval (CI) 1.02–1.28; RD/100,000 doses=0.84, 95% CI 0.14−1.54) compared to the control interval. However, no significant primary analysis results were identified in 2017-2018 or 2018-2019.

**Table 1.**
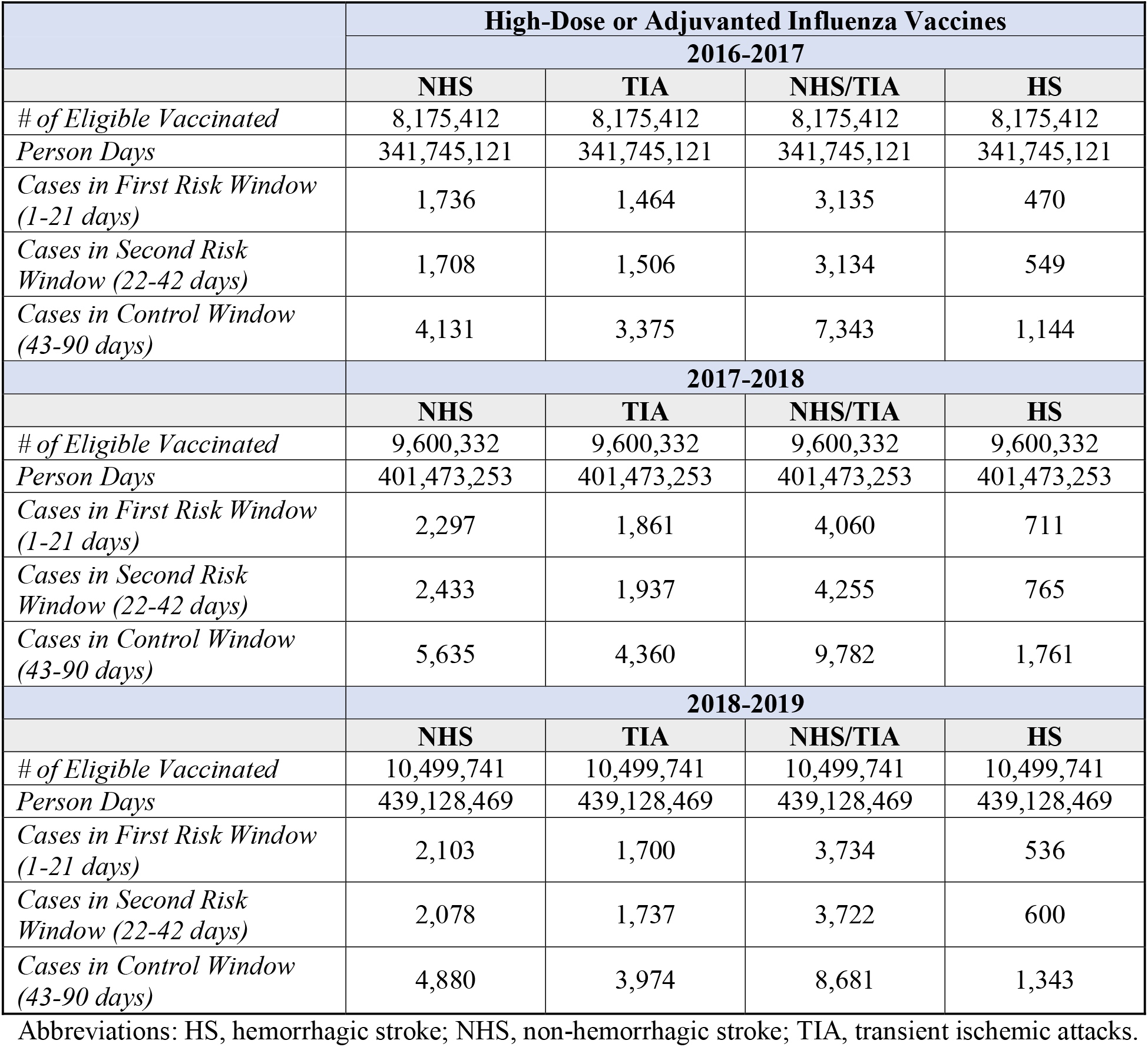
Observed Outcome Counts and Person Days Among Eligible High-Dose or Adjuvanted Influenza Vaccinated Medicare Beneficiaries ≥65 Years, by Influenza Season.

The age subgroup analyses showed significant associations for HS during the 22-42 day risk window in the 65-74 age subgroup in 2016-2017 (IRR=1.24, 95% CI 1.02−1.51; RD/100,000 doses=0.83, 95% CI 0.07−1.59), and in the 75-84 age subgroup in 2017-2018 (IRR=1.19, 95% CI 1.03−1.37; RD/100,000 doses=1.56, 95% CI 0.25−2.87). In 2018-2019 we identified significant associations in the 85+ age subgroup for NHS (IRR=1.17, 95% CI 1.06−1.29; RD/100,000 doses=6.40, 95% CI 2.34−10.47) and NHS/TIA (IRR=1.14, 95% CI 1.06−1.23, RD/100,000 doses=8.66, 95% CI 3.51−13.81) during the 1-21 day risk window.

## Discussion

Overall, we did not observe clear, consistent evidence of increased stroke risk following high-dose or adjuvanted influenza vaccination across the three seasons 2016-2019. The statistically significant associations we identified were not consistently observed across outcomes, risk windows, age subgroups, or seasons. We did not adjust for multiple testing, because the system was designed to be sensitive to potential safety signals. The temporal scan identified case clusters in control windows for several outcomes in 2017-2018 and 2018-2019, which conflicts with the signals identified through the SCCS framework. While studies in the literature show that influenza infections increase the risk of stroke, our study only included vaccinated cases and did not account for the protective effect of vaccination against infections.^2,3^ The clinical significance of any potential risk of stroke following vaccination must be carefully considered together with the significant benefits of receiving an influenza vaccination.^4,5^

## Data Availability

Data produced in the present work are contained in the report. Access to original datasets is not available to protect patient information.

## Acknowledgements

To Henry T. Zhang^1^, PhD for contributions to methodology development, results review, letter writing and review.

To Jeffrey A. Kelman^3^, MD for contributions to methodology development and results review.

To Nathan Duma^2^, PhD; Yiyun Chiang^2^, MPH; and Hai Lyu^2^, MS, for contributions to methodology development, data collection and analysis, and results review.

To Gita Nadimpalli^2^, PhD for contributions to methodology development.

## Author’s contributions

contributed to study design, data collection and analysis, letter writing and review. All authors have seen and approved the manuscript.

## Institutional Review Board (IRB) and Patient Informed Consent

This study did not require full FDA IRB committee review and approval because it was determined to be exempt from the requirements of 45 CFR Part 46; 46.104(d)(2) under the 2018 Common Rule. The use of Medicare administrative data was approved by CMS privacy board under a data use agreement. The analyses utilized only existing records and the subjects cannot be identified. Patient consent was not required since our study was based on CMS claims data and thus does not include factors necessitating patient consent.

## Data Sharing

All data produced in the present work are contained in the manuscript. Access to original datasets is not available to protect patient information.

---

**Figure 1.**
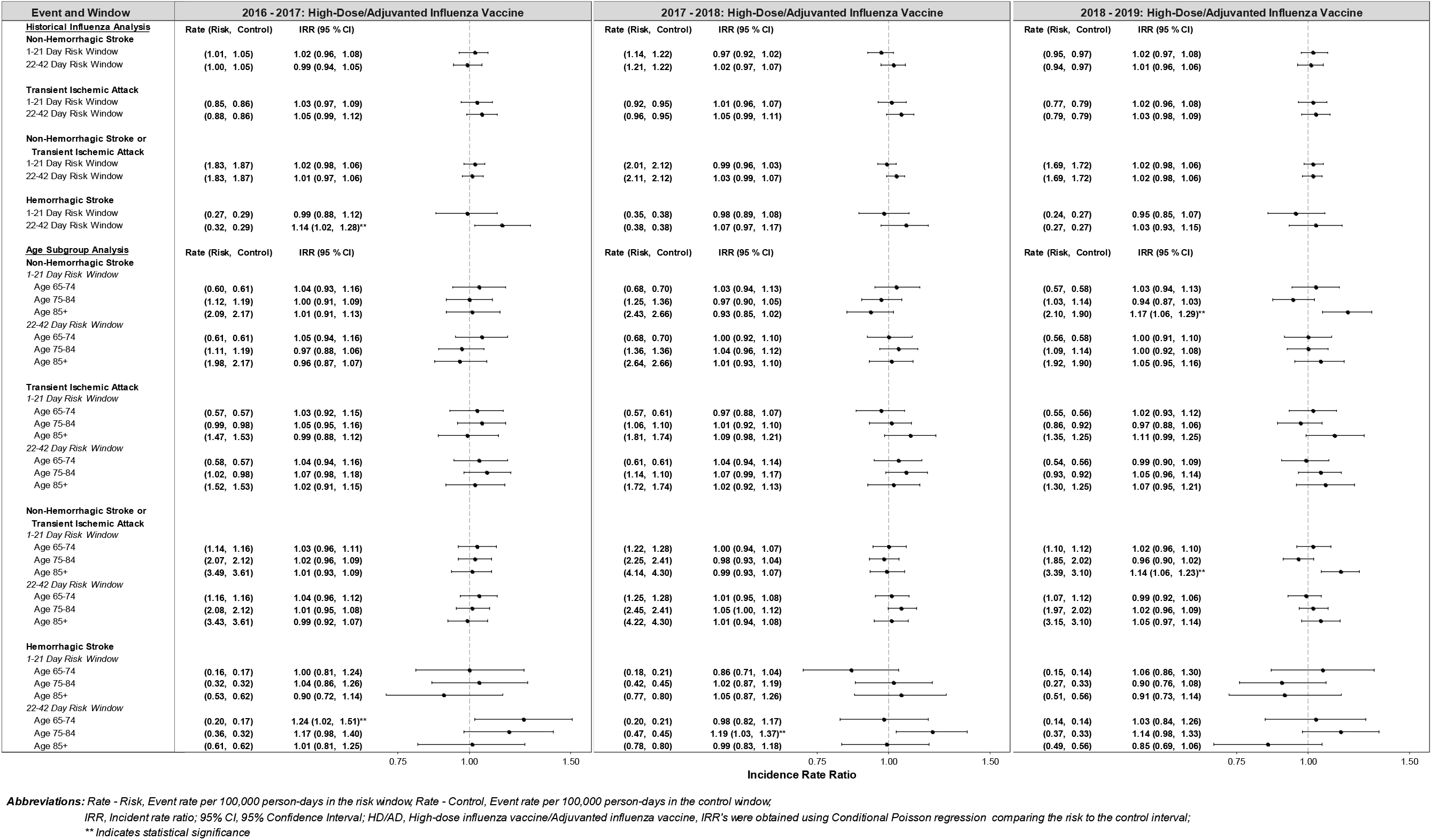
Risk of Stroke Following High-Dose or Adjuvanted Influenza Vaccines: Summary of Primary Analysis and Age Subgroup Analysis by Influenza Season.

